# People with HIV Have Higher Risk of COVID-19 Diagnosis but Similar Outcomes than the General Population

**DOI:** 10.1101/2022.01.13.22269178

**Authors:** Michael E Tang, Thaidra Gaufin, Ryan Anson, Wenhong Zhu, William C Mathews, Edward R Cachay

**Author notes:** **Please address correspondence to:** Edward Cachay MD, MAS, 200 West Arbor Drive, San Diego, CA 92103-8186. **Disclosures:** All authors have completed the ICMJE uniform disclosure form at www.icmje.org/coi_disclosure.pdf and declare: ERC receives unrestricted research funds from Gilead Sciences and Merck Sharp & Dohme for unrelated research, which are paid to UC Regents, ERC also participates in the medical advisory board for Gilead Sciences; no other relationships or activities that could appear to have influenced the submitted work.

## Abstract

**Background:** We investigated the effect of HIV on COVID-19 outcomes with attention to selection bias due to differential testing and to comorbidity burden.

**Methods:** Retrospective cohort analysis using four hierarchical outcomes: positive SARS-CoV-2 test, COVID-19 hospitalization, intensive care unit (ICU) admission, and hospital mortality. The effect of HIV status was assessed using traditional covariate-adjusted, inverse probability weighted (IPW) analysis based on covariate distributions for testing bias (testing IPWs), HIV infection status (HIV IPWs), and combined models. Among PWH, we evaluated whether CD4 count and HIV plasma viral load (pVL) discriminated between those who did or did not develop study outcomes using receiver operating characteristic analysis.

**Results:** Between March and November 2020, 63,319 people were receiving primary care services at UCSD, of whom 4,017 were people living with HIV (PWH). PWH had 2.1 times the odds of a positive SARS-CoV-2 test compared to those without HIV after weighting for potential testing bias, comorbidity burden, and HIV-IPW (95% CI 1.6-2.8). Relative to persons without HIV, PWH did not have an increased rate of COVID-19 hospitalization after controlling for comorbidities and testing bias [adjusted incidence rate ratio (aIRR): 0.5, 95% CI: 0.1 – 1.4]. PWH had neither a different rate of ICU admission (aIRR:1.08, 95% CI; 0.31 – 3.80) nor in-hospital death (aIRR:0.92, 95% CI; 0.08 – 10.94) in any examined model. Neither CD4 count nor pVL predicted any of the hierarchical outcomes among PWH.

**Conclusions:** PWH have a higher risk of COVID-19 diagnosis but similar outcomes compared to those without HIV.

**Summary point:** After considering the effects of potential bias due to differential testing, comorbidities, and other patient characteristics, people with HIV had an increased rate of SARS-CoV-2 positivity and similar rates of hospitalization, ICU admission, and death.

## Introduction

Since the beginning of the COVID-19 pandemic, many have been concerned that people living with HIV (PWH) will have a more severe course of COVID-19 after infection than the general population. Reasons for concern include impaired inflammatory cell responses in PWH, making it more challenging to control respiratory viral infections, leading to more complications and death [1]. PWH are known to have a high prevalence of comorbidities associated with increased COVID-19 disease severity and a high prevalence of barriers to care such as mental illness, drug and alcohol use, and homelessness [1] that impact both risks of acquisition and access to care during an evolving COVID-19 pandemic.

Independent effects of HIV on COVID-19 positivity, hospitalization, and mortality have been inconsistently observed [2–9]. It is unclear whether differences among the studied populations with respect to HIV-induced immunosuppression, prevalence of medical comorbidities, socioeconomic disparities, or methodological issues account for the disparate results. Among methodological considerations are differences in study samples [4,10], variant preponderance [3,11–13], adequacy of control for selection bias [14] and confounding [15]. All observational studies presented have lacked appropriate control for potential collider bias, which can induce a spurious association between the exposure and outcome. We performed a nested cohort study in San Diego, California to evaluate the effect of HIV infection status on hierarchical outcomes of COVID-19 disease. We focused on controlling for comorbidity burden, HIV immunosuppression status, and selection bias due to differential SARS-CoV-2 testing.

## Methods

### Study design and population

The study sample represented a nested sub-cohort of adults (≥18 years) receiving primary care at UC San Diego (UCSD) between March 1, 2020, and November 30, 2020. Receipt of primary care at the study institution was defined as having a primary care provider associated with UCSD and attending at least one primary care appointment from March 1, 2017 to March 1, 2020. We included all adults tested for SARS-CoV-2 during the study period. Patients were classified as SARS-CoV-2 positive as of the date of their first positive test, while those who consistently tested negative were classified SARS-CoV-2 negative. We excluded 28 individuals who received part of their care for any COVID-19 related illness outside UCSD to avoid system differences in management practices (**Supplementary Figure 1**). The study protocol was approved by the UCSD institutional human research protection program (#201226X).

### Study Variables

Study variables were extracted from Clarity database by the Altman Clinical and Translational Research Institute bioinformatics services. Patient characteristics included: A) demographics: age, gender, race/ethnicity; B) conditions associated with immune suppression: history of any organ transplant (solid or bone marrow), active malignancy except basal cell cancer, HIV infection, and rheumatologic disease; C) medical comorbidities according to ICD-10 diagnosis: hypertension (HTN), diabetes mellitus (DM), cardiovascular disease (CVD), coronary artery disease (CAD), congestive heart failure (CHF), chronic pulmonary disease, chronic liver disease, chronic kidney disease (CKD), obesity, mental illness, and the Charlson Comorbidity Index (CCI) [16]; D) use of tobacco, alcohol, recreational drugs, and homelessness. The dates and results were noted for SARS-CoV-2 testing. We also collected HIV transmission risk factors, CD4 cell count, HIV plasma viral load (pVL), and antiretroviral therapy (ART). For HIV time-varying covariates, we collected the most recent value before SARS-CoV-2 testing, but no longer than 12 months before testing. We performed manual chart reviews of everyone diagnosed with COVID-19 to verify SARS-CoV-2 testing and hospital-related outcomes. For HIV-infected subjects, chart review validated values for CD4, pVL, and ART status.

### Study outcomes and statistical analysis

Four hierarchical outcomes were examined among the SARS-CoV-2 tested sub-cohort: SARS-CoV-2 positivity, COVID-19 related hospitalization, intensive care unit (ICU) admission, and hospital mortality. The primary exposure of interest was HIV infection. We estimated the date of SARS-CoV-2 infection as five days before the first positive test date based on the estimated median and mean incubation periods at the time [17,18]. Time-at-risk for the hospital outcomes (hospital admission, ICU admission, hospital death) began with the estimated date of infection and ended on the earliest of: the date of the respective hospital outcome, the last documented health care encounter at the study institution (conservative definition), or the end of the study period (liberal definition). Because conclusions from Poisson regression models did not differ according to censoring definition (conservative or liberal), only results using the liberal definition are presented.

Using contingency table analysis, we evaluated associations between patient characteristics and: (1) being tested for SARS-CoV-2; (2) testing positive for SARS-CoV-2 given testing; (3) hospital outcomes (admission, ICU care, and death) and (4) being HIV-infected. Associations were evaluated using Wilcoxon rank-sum tests for continuous variables and either chi-square or Fisher exact tests for categorical variables. Direction of associations between covariates and testing, positivity given testing, and HIV are presented using odds ratios [95% C.I].

Because of the potential for the selection form of collider bias due to differential testing of those in the source population, inverse probability weights (IPW) of being tested one or more times were calculated based on distributions in the source population [19,20]. IPWs for SARS-CoV-2 testing (testing IPWs) were constructed using logistic regression including age, sex, race, smoking, alcohol, illicit drugs, HTN, DM, CAD, CHF, CVD, pulmonary disease, rheumatic disease, liver disease, CKD, any cancer, solid organ transplant, mental illness diagnosis, peripheral vascular disease, cerebrovascular accident, and CCI. In addition, we constructed IPWs for our primary exposure of interest, HIV infection based on covariate distributions in the source population. HIV-IPWs were calculated using the same covariates used to construct testing IPWs. Further, IPWs were stabilized and 1% truncated using previously described methods [21]. Combined testing and HIV-IPWs were calculated as the product of both contributing weights.

To estimate the effect of HIV infection status on study outcomes, multiple regression analyses were employed. For the outcome SARS-CoV-2 positivity, we fit multiple logistic regression models. For the hospital outcomes (admission, ICU care, mortality), we fit Poisson regression models and estimated incidence rate ratios (IRRs). Because a key covariate (body mass index) was missing in 3.1% of the source health care cohort, multiple imputation incorporating all other non-missing covariates in the imputation model were used. The effect of HIV status on study outcomes was estimated using the following estimation approaches: (1) unadjusted and unweighted; (2) unadjusted with testing IPWs applied; (3) unadjusted with HIV-IPWs applied; (4) unadjusted with combined IPWs applied; (5) unweighted traditional covariate adjustment; and (6) traditional covariate adjustment with testing IPWs applied. Traditional covariate-adjusted models included those covariates found to be significant predictors of SARS-CoV-2 positivity in bivariate analysis. Reported confidence intervals were based on robust standard errors. To assess the potential impact of residual confounding of the effect of HIV status on study outcomes, we calculated E-values, which are defined as the minimum strength of association on the effect measure scale that an unmeasured confounder would need to have with both HIV status and the outcome to fully explain away a specific exposure-outcome association, conditional on the measured covariates [22,23].

We investigated whether CD4 cell count and pVL discriminated between those who developed or did not develop study outcomes using receiver operating characteristic (ROC) analysis. We assigned each study outcome as a criterion measure evaluated against absolute CD4 count and log10-transformed pVL as classification measures.

Data were stored in REDCap (Research Electronic Data Capture, Vanderbilt University, Nashville, TN). All tests were two-tailed, using α = 0.05 and performed using Stata version 17.0.

## Results

Between March 1 to November 30, 2020, 63,319 people were receiving primary care services at UCSD Health, of whom 4,017 were PWH. People who were older, white, obese, or who had any significant comorbidity, HIV infection, mental illness, drug use, or homelessness were more likely to be tested for SARS-CoV-2 than those without those characteristics (**Supplementary Table 1**). Compared to HIV-uninfected patients ever tested for SARS-CoV-2, tested PWH were more commonly younger, male, obese, and non-white. PWH were also more likely to have documented smoking, recreational drug use, homelessness, and to have diagnoses of HTN, DM, myocardial infarction, liver disease, chronic pulmonary disease, and mental illness (**Supplementary Table 2**). During the study period, 487 patients tested positive for SARS-CoV-2, including 88 PWH. Of the 88 PWH diagnosed with COVID-19, 6 were hospitalized for COVID-19 related complications, 4 were admitted to the ICU, and 1 died in the hospital. In contrast, among 399 HIV-uninfected patients diagnosed with COVID-19, there were 57 admitted to the hospital, 19 admitted to the ICU and 7 died in the hospital. In bivariate analysis of those ever tested for SARS-CoV-2, persons with a positive SARS-CoV-2 test were more commonly HIV-infected, obese, homeless, and using recreational drugs. Several covariates were associated with discordance between SARS-CoV-2 testing and test results given testing. Non-white and younger people were less likely to be tested but were more likely to have a positive SARS-CoV-2 result if tested (**Supplementary Table 3**). Although patients with HTN, CVD, solid cancer, and higher comorbidity burden by CCI were more often tested, they were less likely to test positive for SARS-CoV-2 (**Table 1**).

**Table 1:** Unadjusted Odds Ratios with 95% confidence intervals for ever-tested, HIV+ and any positive SARS-CoV-2 patients

| Independent Variable | Dependent Variable |  |  |
| --- | --- | --- | --- |
|  | Ever-tested<br>(n=62319) | HIV<br>(n=62319) | Any positive<br>(n=16626) |
| **HIV** | 1.222***<br>[1.140, 1.310] |  | 2.910***<br>[2.292, 3.694] |
| **AGE ≥ 50 years old** | 1.467***<br>[1.415, 1.521] | 0.535***<br>[0.501, 0.572] | 0.381***<br>[0.316, 0.460] |
| **Male Sex** | 1.009<br>[0.974, 1.046] | 0.119***<br>[0.109, 0.130] | 0.757**<br>[0.632, 0.906] |
| **Non-white race** | 0.785***<br>[0.756, 0.815] | 1.911***<br>[1.792, 2.038] | 2.112***<br>[1.763, 2.530] |
| **Smoking** | 0.840***<br>[0.778, 0.906] | 6.996***<br>[6.459, 7.578] | 0.732<br>[0.466, 1.151] |
| **Hypertension** | 1.382***<br>[1.328, 1.438] | 1.132***<br>[1.053, 1.217] | 0.778*<br>[0.632, 0.959] |
| **Any diabetes mellitus** | 1.358***<br>[1.281, 1.441] | 1.047<br>[0.938, 1.169] | 1.155<br>[0.878, 1.521] |
| **Cardiovascular disease** | 1.752***<br>[1.661, 1.849] | 1.039<br>[0.938, 1.152] | 0.642**<br>[0.476, 0.865] |
| **Homelessness** | 1<br>[1, 1] | 29.41***<br>[22.41, 38.58] | 1.476<br>[0.778, 2.799] |
| **Alcohol use** | 1.004<br>[0.969, 1.040] | 0.479***<br>[0.449, 0.512] | 0.933<br>[0.778, 1.118] |
| **Illicit drugs use** | 1.256***<br>[1.180, 1.337] | 4.782***<br>[4.426, 5.166] | 1.333*<br>[1.010, 1.758] |
| **Myocardial infarction** | 1.882***<br>[1.606, 2.206] | 2.095***<br>[1.654, 2.654] | 0.919<br>[0.431, 1.959] |
| **Congestive heart failure** | 1.865***<br>[1.702, 2.043] | 1.029<br>[0.860, 1.230] | 0.707<br>[0.434, 1.154] |
| **Peripheral vascular disease** | 8.516***<br>[5.319, 13.63] | 1.168<br>[0.540, 2.525] | 1<br>[1, 1] |
| **Cerebrovascular accident** | 1.522***<br>[1.395, 1.660] | 1.140<br>[0.971, 1.338] | 0.694<br>[0.425, 1.131] |
| **Dementia** | 0.900<br>[0.771, 1.050] | 0.671*<br>[0.486, 0.926]" | 0.305<br>[0.0757, 1.233] |
| **Pulmonary disease** | 1.634***<br>[1.541, 1.733]" | 1.619***<br>[1.469, 1.784] | 0.782<br>[0.575, 1.064] |
| **Rheumatological disease** | 1.770***<br>[1.554, 2.015] | 0.178***<br>[0.101, 0.315] | 0.526<br>[0.234, 1.184] |
| **Mild liver disease** | 1.655***<br>[1.540, 1.779] | 2.882***<br>[2.609, 3.183] | 1.297<br>[0.952, 1.768] |
| **Severe liver disease** | 2.486***<br>[1.969, 3.138] | 2.668*** | 0.246<br>[0.0343, 1.761] |
| **Hemiplegia** | 1.281<br>[0.960, 1.709] | 2.854***<br>[1.984, 4.106] | 1.004<br>[0.245, 4.112] |
| **Chronic kidney disease** | 1.619***<br>[1.508, 1.739] | 1.369***<br>[1.210, 1.549] | 1.170<br>[0.851, 1.608] |
| **Lymphoma** | 2.041***<br>[1.714, 2.430] | 2.380***<br>[1.854, 3.054] | 0.605<br>[0.224, 1.633] |
| **leukemia** | 2.263***<br>[1.918, 2.671] | 0.334***<br>[0.193, 0.580] | 1.061<br>[0.522, 2.159] |
| **Solid cancer** | 1.961***<br>[1.852, 2.076] | 0.884*<br>[0.786, 0.995] | 0.467***<br>[0.327, 0.666] |
| **Metastatic cancer** | 2.224***<br>[1.962, 2.521] | 0.275***<br>[0.174, 0.433] | 0.596<br>[0.294, 1.206] |
| **Obesity (BMI≥ 30)** | 1.074***<br>[1.030, 1.119] | 1.111**<br>[1.032, 1.198] | 1.579***<br>[1.303, 1.914] |
| **Mental illness** | 1.394***<br>[1.342, 1.447] | 4.571***<br>[4.276, 4.886] | 0.973<br>[0.805, 1.177] |
| **Solid organ transplant** | 2.876***<br>[2.417, 3.421] | 0.433**<br>[0.259, 0.725] | 1.322<br>[0.698, 2.503] |
| **Bone Marrow transplant** | 2.655***<br>[2.022, 3.486] | 0.430*<br>[0.191, 0.970] | 1.004<br>[0.317, 3.178] |
| **Charlson comorbidity index** | 1.125***<br>[1.117, 1.133] | 0.904***<br>[0.890, 0.919] | 0.840***<br>[0.803, 0.879] |

Exponentiated coefficients; 95% confidence intervals in brackets
\* p<0.05
\*\* p<0.01
\*\*\* p<0.001

### HIV and SARS-CoV-2 Positivity

PWH had greater odds of testing positive for SARS-CoV-2 than patients without HIV in all logistic regression models (**Table 2**). PWH had 2.9 times the odds of having a positive SARS-CoV-2 result in unadjusted analysis (95% CI 2.3-3.7). After testing IPWs, PWH still had 2.8 times the odds of testing positive for SARS-CoV-2 compared to patients without HIV. When HIV-IPWs were used, PWH had 3.5 times the odds for testing positive than patients without HIV (95% CI 2.6-4.9). This effect was similar when combining testing IPWs and HIV-IPWs. However, the impact of HIV on SARS-CoV-2 positivity decreased in traditional comorbidity covariate adjustment to having 2.2 times the odds (95% CI 1.7-2.8). Finally, when traditional covariate adjustment was combined with testing IPWs, PWH still had a 2.1 times the odds for testing positive for SARS-CoV-2 compared to patients without HIV (95% CI 1.6-2.8).

**Table 2:** Effects of HIV on COVID-19 status, hospitalization, ICU admission and in-hospital death in poisson regression models (unadjusted, inverse probability weighted, adjusted and combined)

|  | (1)<br>Unadjusted &<br>unweighted | (2)<br>Testing IPW | (3)<br>IPW:HIV | (4)<br>IPW:Combined | (5)<br>Covariate-Adjusted | (6)<br>Covariate-Adjusted &<br>Testing IPW |
| --- | --- | --- | --- | --- | --- | --- |
| **Outcome:** |  |  |  |  |  |  |
| **Any positive SARS-CoV-2 test** |  |  |  |  |  |  |
| Odds Ratio (HIV+) | 2.910*** | 2.834*** | 3.547*** | 3.407*** | 2.153*** | 2.121*** |
| 95% Confidence Interval | [2.292–3.694] | [2.216–3.624] | [2.576–4.884] | [2.469–4.700] | [1.664–2.787] | [1.619–2.778] |
| Observations | 16626 | 16625 | 16625 | 16625 | 16625 | 16625 |
| E-value | 5.268 | 5.113 | 6.552 | 6.27 | 3.8 | 3.762 |
| **Outcome: Hospitalization** |  |  |  |  |  |  |
| Incidence Rate Ratios (HIV+) | 0.316* | 0.449 | 0.234* | 0.362 | 0.338* | 0.482 |
| 95% Confidence Interval | [0.127–0.788] | [0.169–1.191] | [0.068–0.802] | [0.103–1.281] | [0.132–0.865] | [0.169–1.375] |
| E-value | 5.783 | 3.882 | 8.008 | 4.961 | 5.629 | 3.731 |
| Observations | 486 | 485 | 485 | 485 | 485 | 485 |
| **Outcome: ICU admission** |  |  |  |  |  |  |
| Incidence Rate Ratios (HIV+) | 0.764 | 0.928 | 0.533 | 0.793 | 0.924 | 1.084 |
| 95% Confidence Interval | [0.260–2.247] | [0.296–2.910] | [0.133–2.141] | [0.175–3.604] | [0.298–2.860] | [0.310–3.800] |
| E-value | 1.943 | 1.367 | 3.155 | 1.834 | 1.55 | 1.211 |
| Observations | 487 | 486 | 486 | 486 | 486 | 486 |
| **Outcome: Hospital Death** |  |  |  |  |  |  |
| Incidence Rate Ratios (HIV+) | 0.519 | 0.583 | 0.329 | 0.372 | 0.729 | 0.924 |
| 95% Confidence Interval | [0.064 - 4.214] | [0.070 - 4.831] | [0.039 - 2.745] | [0.044 - 3.123] | [0.078 - 6.813] | [0.078 - 10.943] |
| E-value | 3.267 | 2.824 | 5.538 | 4.82 | 2.084 | 1.381 |
| Observations | 487 | 486 | 486 | 486 | 486 | 486 |

1. IPWs standardized for age, sex, race, smoking, alcohol, illicit drugs, hypertension, diabetes mellitus, coronary artery disease, congestive heart failure, cardiovascular disease, pulmonary disease, rheumatic disease, liver disease, chronic kidney disease, any cancer, solid organ transplant, mental illness diagnosis, peripheral vascular disease, cerebrovascular accident & Charlson score. 2. Covariate-adjusted estimates included covariates: obesity, age, sex, race, hypertension, diabetes mellitus, cardiovascular disease, solid cancer & Charlson comorbidity index.
\* p<0.05; \*\* p<0.01; \*\*\* p<0.001

### HIV and rate of COVID-19 hospitalization

The median days from estimated date of infection to hospitalization for PWH was 5.5 days (IQR 1-13 days) compared with 6 days (IQR 5-11 days) for patients without HIV (Wilcoxon rank sum p = 0.46). Yet, the probability of hospitalization following a positive SARS-CoV-2 test appeared lower for PWH than patients without HIV (**Figure 1**). In unadjusted and unweighted analysis, PWH were 70% less likely to be admitted to the hospital for COVID-19 than patients without HIV (IRR 0.3 [95% CI 0.1-0.8]). PWH remained less likely to be admitted to the hospital for COVID-19 compared to patients without HIV when traditional covariate adjustment was performed (aIRR 0.3 [95% CI 0.1-0.9]) and when HIV-IPWs are applied (IRR 0.2 [95% CI 0.1-0.8]). In other models of hospitalization, effect measures were similar in direction and magnitude but not statistically significant at conventional levels (**Table 2**).

**Figure 1:**
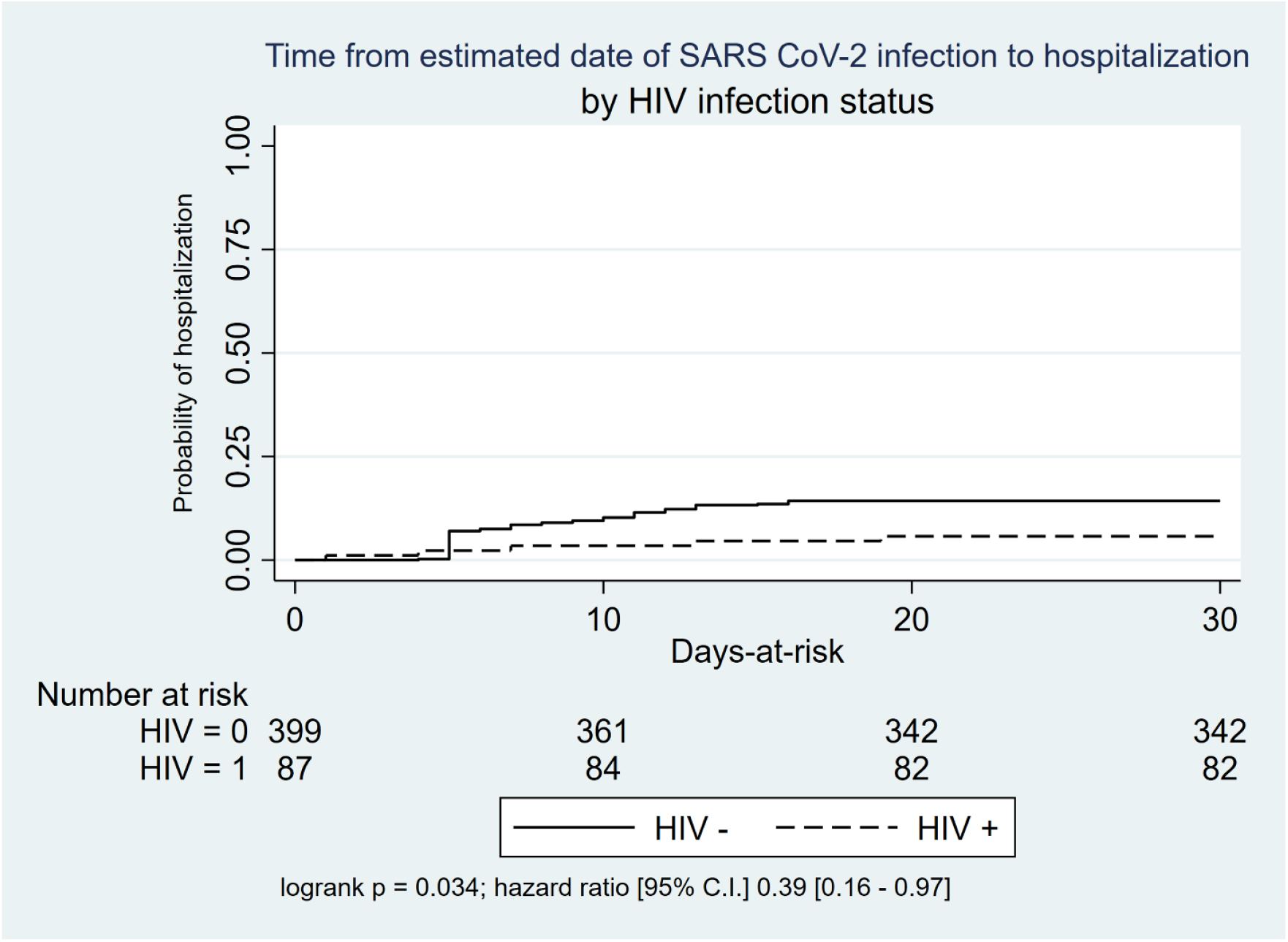
Kaplan Meier curve for the time from estimated date of SARS-CoV-2 infection to hospitalization by HIV infection status

### HIV and risk of ICU admission

There was no difference in the rate of ICU admission among PWH and patients without HIV diagnosed with COVID-19, with an unadjusted IRR of 0.8 (95% CI 0.3-2.2) (**Table 2**). After applying testing IPWs alone, HIV-IPWs alone, and combined IPWs, there was still no difference in the likelihood of ICU admission by HIV status (IRR 0.9 [95% CI 0.3-2.9], 0.5 [95% CI 0.1-2.1], and 0.8 [95% CI 0.2-3.6] respectively). Traditional covariate adjustment found no difference in the rate of ICU admission between PWH and patients without HIV, both with testing IPWs (aIRR 1.1 [95% CI 0.3-3.8]) and without (aIRR 0.9 [95% CI 0.3-2.9]).

### HIV and risk of in-hospital death

There was no difference in hospital death rates among PWH and patients without HIV diagnosed with COVID-19, with an unadjusted IRR of 0.5 (95% CI 0.1-4.2) (**Table 2**). These findings did not change with the inclusion of testing IPWs, HIV-IPWs, traditional covariate adjustment, or the combination of covariate adjustment with testing IPWs.

### CD4 cell count and HIV viral load and COVID-19 outcomes

The median [interquartile range] of absolute CD4 counts among PWH in the source population (n=1126) and among the COVID-19 positive PWH (n=84) were 634 [405, 881] and 638 [407, 1000], respectively. For pVL, 85% and 88% were undetectable (≤50 copies/ml) in the source population and in the COVID-19 positive PWH, respectively. In ROC analysis, neither CD4 count nor pVL discriminated between PWH who tested positive for SARS-CoV-2, PWH who were admitted to the hospital, or PWH who were admitted to the ICU, compared to those without each of these outcomes, respectively. For SARS-CoV-2 positivity, ROC was 0.5 for CD4 count and pVL. For hospitalization, ROC was 0.4 for CD4 count and pVL. For ICU admission, ROC was 0.2 (95% CI 0-0.5) for CD4 count and 0.5 (95% CI 0.3-0.7) for pVL (**Table 3**).

**Table 3:** Receiver Operating Charactistic (ROC) curve analyses for CD4 and log10 HIV viral load discriminant ability for SARS-CoV-2 positivity, COVID-19-related hospitalization or ICU admission among patients with HIV

| Outcome | N | ROC | Standard error | 95% confidence interval | P-value* |
| --- | --- | --- | --- | --- | --- |
| **Any positive SARS-CoV-2 test** |  |  |  |  |  |
| CD4 | 1,104 | 0.55 | 0.04 | 0.48–0.61 | 0.17 |
| HIV viral load | 1,104 | 0.48 | 0.02 | 0.43–0.53 |  |
| **Hospitalization** |  |  |  |  |  |
| CD4 | 81 | 0.35 | 0.11 | 0.13–0.58 | 0.53 |
| HIV viral load | 81 | 0.45 | 0.07 | 0.31–0.59 |  |
| **ICU admission** |  |  |  |  |  |
| CD4 | 81 | 0.26 | 0.13 | 0.01–0.51 | 0.23 |
| HIV viral load | 81 | 0.49 | 0.10 | 0.29–0.69 |  |

\* Compares the null hypothesis that the area under the curve for CD4 cell count is equal to the area under the curve for log10 HIV plasma viral load using chi-square test of significance.

## Discussion

This study found that in 2020, before the widespread availability of COVID-19 vaccines, PWH had more than twice the risk of testing positive for SARS-CoV-2 compared to those without HIV infection. Our study used a nested cohort analysis because, unlike other studies, we access prognostic characteristics of the source population under care that could have been SARS-CoV-2 testing. This approach allowed at least partial control for differential testing by applying testing IPWs. Conclusions regarding excess SARS-CoV-2 positivity among PWH were robust to different modeling approaches.

Despite regional variation in SARS-CoV-2 testing availability and indication, PWH have generally experienced lower thresholds for diagnostic testing [11]. For example, a study from Kaiser Permanente Southern California (KPSC) found a higher incidence of testing, diagnosis, and COVID-19 hospitalization in PWH than in HIV-uninfected patients [24]. Yet, PWH were designated as a potential risk group at KPSC during the study period and were prioritized for testing. Testing threshold as a function of symptom severity, comorbidity, and test availability may contribute to differing observed disease trajectories, including hospital-related outcomes.

A robust conclusion from our analyses is that HIV infection was not associated with increased COVID-19 hospitalization rate. Like other studies, HIV status did not affect ICU admission or death in any models [6,25]. Our results contrast with studies that observed an increased risk of hospitalization among PWH [6,12,24,26]. However, most did not fully account for differences in comorbidities that likely impact the risk of COVID-19 outcomes [27,28]. When examining COVID-19 outcomes among patients diagnosed with COVID-19, several studies found that PWH have an increased risk of hospitalization but not hospital mortality despite matching for key cofounders including comorbidity burden [6,29]. There are several threats to the validity of conclusions from observational studies of the independent effect of HIV infection on risk for SARS-CoV-2 infection and its hospital-related outcomes. Perhaps the most important are selection bias due to differential testing in defining the study sample, inadequate control for confounding, and missing information regarding the severity of illness and prognostic factors at the time of diagnosis. Our study attempted to control for differential testing and confounding. However, we had limited access to markers of severity of illness at the time of diagnosis of SARS-CoV-2 infection. In particular, because the availability of diagnostic testing was limited and testing thresholds were changing during the study period, the proportion of undiagnosed SARS-CoV-2 infections among either PWH or those without HIV infection is unknown.

As with others studies [7,26,30,31], neither most recent CD4 count nor pVL were discriminators of the risk for COVID-19 diagnosis or hospitalization with or without ICU admission. However, we suggest cautious interpretation concerning the effect of pVL because of range restriction (84% undetectable). One study observed that patients with CD4 count < 200 cells/mm^3^ and those with detectable viremia have a higher risk of severe COVID-19, but it did not adjust for the presence of comorbidities that modify the COVID-19 disease course [27].

Our study is subject to limitations. First, like all observational SARS-CoV-2 studies, we could not implement routine universal SARS-CoV-2 screening to identify all patients with SARS-CoV-2 infection. This limitation is problematic especially for prognostic analyses, because of uncertainty regarding the proportion of included patients being asymptomatic at diagnosis and their COVID-related symptom burden at the time of diagnosis. The observation that median days to hospitalization after estimated date of infection did not differ significantly by HIV status provides some support that trajectories of disease progression were similar for both groups. Second, to reduce bias in our analyses due to missing records, varying standards of care, and referral indication, individuals were excluded if part of the COVID-19 care cascade occurred outside of UCSD Health. The number of excluded patients is unlikely to alter the conclusions based on included patients. Third, conclusions regarding the effect of HIV infection on ICU admission and hospital mortality should be interpreted cautiously because of the small number of ICU and mortality events in the study sample. Fourth, our PWH cohort generally had well-controlled HIV infection, with only 7% of those tested having AIDS and 9% a pVL above 200 copies/mL (**Supplementary Table 4**). Hence, our findings cannot be generalized to PWH populations with higher proportions of unsuppressed viremia. Similarly, we did not assess nadir CD4 counts, which some have suggested is a more reliable marker of immunosuppression risk for severe COVID-19 among PWH [32]. Fifth, although we took into account comprehensive comorbidity measures in estimating effects, residual sources of bias and confounding may remain. E-values for residual confounding (**Table 2**) suggest bounds to assess conditions required to nullify observed effects. Lastly, our use of median of 5 days from infection event to first positive test is subject to several limitations: (1) testing threshold changed over the study period according to availability of tests and index of clinical suspicion; (2) SARS-CoV-2 variant evolution could alter incubation periods, and (3) rates of progression to clinical detection may have been differential with respect to HIV infection status and other covariates [17,18,33].

In conclusion, after considering the effects of differential testing using inverse probability of selection weighting, comorbidity burden, and other patient characteristics, PWH, compared to those without HIV, had an increased rate of SARS-CoV-2 positivity, similar or perhaps a lower hospitalization rate, and similar rates of ICU admission and death.

## Data Availability

All data produced in the present study are available upon reasonable request to the authors

## Acknowledgments

We gratefully acknowledge Margaret Quattrocchi for support in data entry for the HIV COVID-19 registry.

---

**Supplementary Table 1.**
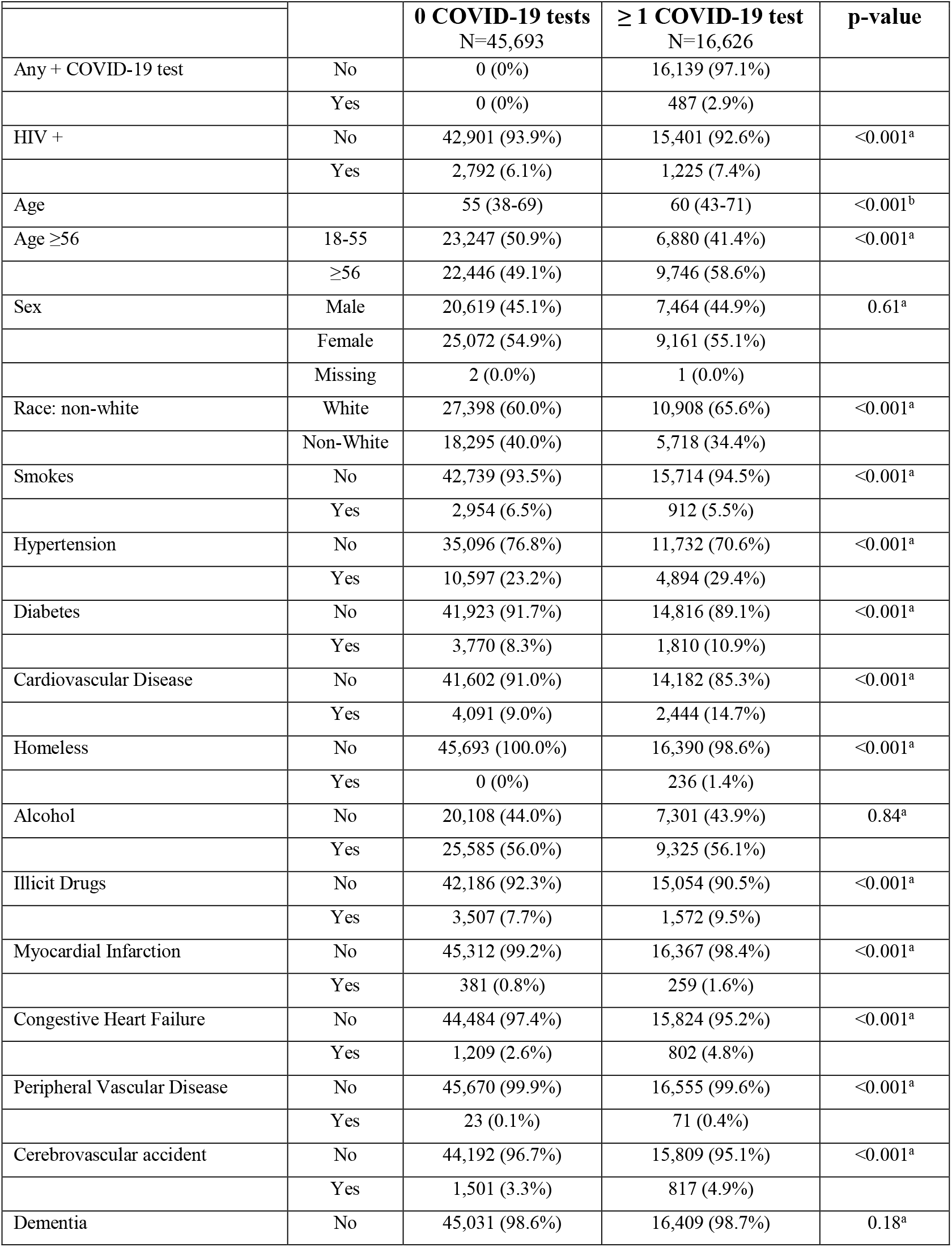
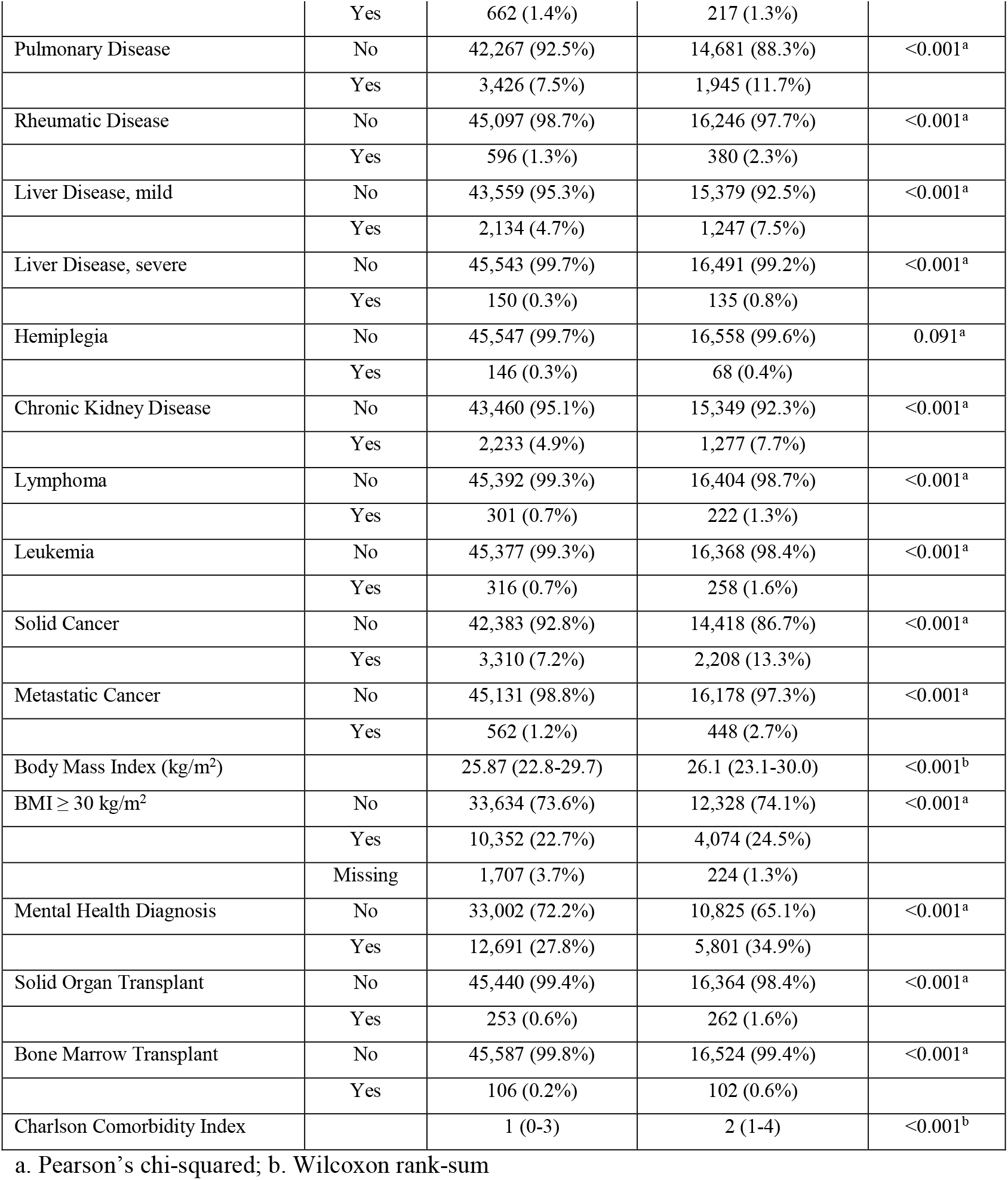
Association between testing for COVID-19 and patient characteristics among all under care at UCSD Health.

**Supplementary Table 2:** Association between HIV infection status and patient characteristics among those ever tested for SARS-CoV-2 at UC San Diego Health

|  |  | **HIV Negative**<br>N=15,401 | **HIV Positive**<br>N=1,225 | **p-value** |
| --- | --- | --- | --- | --- |
| Any + COVID test | No | 15,002 (97.4%) | 1,137 (92.8%) | <0.001<sup>a</sup> |
|  | Yes | 399 (2.6%) | 88 (7.2%) |  |
| Age |  | 61 (44-72) | 54 (43-60) | <0.001<sup>b</sup> |
| Age ≥56 | 18-55 | 6,186 (40.2%) | 694 (56.7%) | <0.001<sup>a</sup> |
|  | ≥56 | 9,215 (59.8%) | 531 (43.3%) |  |
| Sex | Male | 6,406 (41.6%) | 1,058 (86.4%) | <0.001<sup>a</sup> |
|  | Female | 8,994 (58.4%) | 167 (13.6%) |  |
|  | Missing | 1 (0.0%) | 0 (0.0%) |  |
| Race: non-white | White | 10,312 (67.0%) | 596 (48.7%) | <0.001<sup>a</sup> |
|  | Non-White | 5,089 (33.0%) | 629 (51.3%) |  |
| Smokes | No | 14,789 (96.0%) | 925 (75.5%) | <0.001<sup>a</sup> |
|  | Yes | 612 (4.0%) | 300 (24.5%) |  |
| Hypertension | No | 10,908 (70.8%) | 824 (67.3%) | 0.008<sup>a</sup> |
|  | Yes | 4,493 (29.2%) | 401 (32.7%) |  |
| Diabetes | No | 13,739 (89.2%) | 1,077 (87.9%) | 0.16<sup>a</sup> |
|  | Yes | 1,662 (10.8%) | 148 (12.1%) |  |
| Cardiovascular Disease | No | 13,137 (85.3%) | 1,045 (85.3%) | 1.00<sup>a</sup> |
|  | Yes | 2,264 (14.7%) | 180 (14.7%) |  |
| Homeless | No | 15,321 (99.5%) | 1,069 (87.3%) | <0.001<sup>a</sup> |
|  | Yes | 80 (0.5%) | 156 (12.7%) |  |
| Alcohol | No | 6,567 (42.6%) | 734 (59.9%) | <0.001<sup>a</sup> |
|  | Yes | 8,834 (57.4%) | 491 (40.1%) |  |
| Illicit Drugs | No | 14,196 (92.2%) | 858 (70.0%) | <0.001<sup>a</sup> |
|  | Yes | 1,205 (7.8%) | 367 (30.0%) |  |
| Myocardial Infarction | No | 15,173 (98.5%) | 1,194 (97.5%) | 0.004<sup>a</sup> |
|  | Yes | 228 (1.5%) | 31 (2.5%) |  |
| Congestive Heart Failure | No | 14,658 (95.2%) | 1,166 (95.2%) | 0.99<sup>a</sup> |
|  | Yes | 743 (4.8%) | 59 (4.8%) |  |
| Peripheral Vascular Disease | No | 15,335 (99.6%) | 1,220 (99.6%) | 0.92<sup>a</sup> |
|  | Yes | 66 (0.4%) | 5 (0.4%) |  |
| Cerebrovascular Accident | No | 14,649 (95.1%) | 1,160 (94.7%) | 0.51<sup>a</sup> |
|  | Yes | 752 (4.9%) | 65 (5.3%) |  |
| Dementia | No | 15,198 (98.7%) | 1,211 (98.9%) | 0.60<sup>a</sup> |
|  | Yes | 203 (1.3%) | 14 (1.1%) |  |
| Pulmonary Disease | No | 13,658 (88.7%) | 1,023 (83.5%) | <0.001<sup>a</sup> |
|  | Yes | 1,743 (11.3%) | 202 (16.5%) |  |
| Rheumatic Disease | No | 15,026 (97.6%) | 1,220 (99.6%) | <0.001<sup>a</sup> |
|  | Yes | 375 (2.4%) | 5 (0.4%) |  |
| Liver Disease, mild | No | 14,364 (93.3%) | 1,015 (82.9%) | <0.001<sup>a</sup> |
|  | Yes | 1,037 (6.7%) | 210 (17.1%) |  |
| Liver Disease, severe | No | 15,285 (99.2%) | 1,206 (98.4%) | 0.003<sup>a</sup> |
|  | Yes | 116 (0.8%) | 19 (1.6%) |  |
| Hemiplegia | No | 15,339 (99.6%) | 1,219 (99.5%) | 0.65<sup>a</sup> |
|  | Yes | 62 (0.4%) | 6 (0.5%) |  |
| Chronic Kidney Disease | No | 14,254 (92.6%) | 1,095 (89.4%) | <0.001<sup>a</sup> |
|  | Yes | 1,147 (7.4%) | 130 (10.6%) |  |
| Lymphoma | No | 15,201 (98.7%) | 1,203 (98.2%) | 0.14<sup>a</sup> |
|  | Yes | 200 (1.3%) | 22 (1.8%) |  |
| Leukemia | No | 15,150 (98.4%) | 1,218 (99.4%) | 0.004<sup>a</sup> |
|  | Yes | 251 (1.6%) | 7 (0.6%) |  |
| Solid Cancer | No | 13,328 (86.5%) | 1,090 (89.0%) | 0.015<sup>a</sup> |
|  | Yes | 2,073 (13.5%) | 135 (11.0%) |  |
| Metastatic Cancer | No | 14,958 (97.1%) | 1,220 (99.6%) | <0.001<sup>a</sup> |
|  | Yes | 443 (2.9%) | 5 (0.4%) |  |
| Body Mass Index (kg/m<sup>2</sup>) |  | 26.1 (23.0-30.0) | 26.9 (24.0-30.4) | <0.001<sup>b</sup> |
| BMI ≥ 30 kg/m<sup>2</sup> | No | 11,453 (74.4%) | 875 (71.4%) | 0.074<sup>a</sup> |
|  | Yes | 3,741 (24.3%) | 333 (27.2%) |  |
|  | Missing | 207 (1.3%) | 17 (1.4%) |  |
| Mental Health Diagnosis | No | 10,467 (68.0%) | 358 (29.2%) | <0.001<sup>a</sup> |
|  | Yes | 4,934 (32.0%) | 867 (70.8%) |  |
| Solid Organ Transplant | No | 15,148 (98.4%) | 1,216 (99.3%) | 0.014<sup>a</sup> |
|  | Yes | 253 (1.6%) | 9 (0.7%) |  |
| Bone Marrow Transplant | No | 15,301 (99.4%) | 1,223 (99.8%) | 0.036<sup>a</sup> |
|  | Yes | 100 (0.6%) | 2 (0.2%) |  |
| Charlson Comorbidity Index |  | 2 (1-4) | 2 (0-3) | <0.001<sup>b</sup> |

a. Pearson's chi-squared; b. Wilcoxon rank-sum

**Supplementary Table 3:** Association between any positive SARS-CoV-2 test result and patient characteristics among those ever tested

|  |  | **SARS-CoV-2 Negative** | **SARS-CoV-2 Positive** | **p-value** |
| --- | --- | --- | --- | --- |
|  |  | N=16,139 | N=487 |  |
| HIV + | No | 15,002 (93.0%) | 399 (81.9%) | <0.001<sup>a</sup> |
|  | Yes | 1,137 (7.0%) | 88 (18.1%) |  |
| Age |  | 60 (44-71) | 48 (35-62) | <0.001<sup>b</sup> |
| Age ≥56 | 18-55 | 6,567 (40.7%) | 313 (64.3%) | <0.001<sup>a</sup> |
|  | ≥56 | 9,572 (59.3%) | 174 (35.7%) |  |
| Sex | Male | 7,213 (44.7%) | 251 (51.5%) | 0.002<sup>a</sup> |
|  | Female | 8,926 (55.3%) | 235 (48.3%) |  |
|  | Missing | 0 (0.0%) | 1 (0.2%) |  |
| Race: non-white | White | 10,674 (66.1%) | 234 (48.0%) | <0.001<sup>a</sup> |
|  | Non-White | 5,465 (33.9%) | 253 (52.0%) |  |
| Smokes | No | 15,247 (94.5%) | 467 (95.9%) | 0.18<sup>a</sup> |
|  | Yes | 892 (5.5%) | 20 (4.1%) |  |
| Hypertension | No | 11,365 (70.4%) | 367 (75.4%) | 0.018<sup>a</sup> |
|  | Yes | 4,774 (29.6%) | 120 (24.6%) |  |
| Diabetes | No | 14,389 (89.2%) | 427 (87.7%) | 0.30<sup>a</sup> |
|  | Yes | 1,750 (10.8%) | 60 (12.3%) |  |
| Cardiovascular Disease | No | 13,744 (85.2%) | 438 (89.9%) | 0.003<sup>a</sup> |
|  | Yes | 2,395 (14.8%) | 49 (10.1%) |  |
| Homeless | No | 15,913 (98.6%) | 477 (97.9%) | 0.23<sup>a</sup> |
|  | Yes | 226 (1.4%) | 10 (2.1%) |  |
| Alcohol | No | 7,079 (43.9%) | 222 (45.6%) | 0.45<sup>a</sup> |
|  | Yes | 9,060 (56.1%) | 265 (54.4%) |  |
| Illicit Drugs | No | 14,626 (90.6%) | 428 (87.9%) | 0.042<sup>a</sup> |
|  | Yes | 1,513 (9.4%) | 59 (12.1%) |  |
| Myocardial Infarction | No | 15,887 (98.4%) | 480 (98.6%) | 0.83<sup>a</sup> |
|  | Yes | 252 (1.6%) | 7 (1.4%) |  |
| Congestive Heart Failure | No | 15,354 (95.1%) | 470 (96.5%) | 0.16<sup>a</sup> |
|  | Yes | 785 (4.9%) | 17 (3.5%) |  |
| Peripheral Vascular Disease | No | 16,068 (99.6%) | 487 (100.0%) | 0.14<sup>a</sup> |
|  | Yes | 71 (0.4%) | 0 (0.0%) |  |
| Cerebrovascular Accident | No | 15,339 (95.0%) | 470 (96.5%) | 0.14<sup>a</sup> |
|  | Yes | 800 (5.0%) | 17 (3.5%) |  |
| Dementia | No | 15,924 (98.7%) | 485 (99.6%) | 0.078<sup>a</sup> |
|  | Yes | 215 (1.3%) | 2 (0.4%) |  |
| Pulmonary Disease | No | 14,240 (88.2%) | 441 (90.6%) | 0.12<sup>a</sup> |
|  | Yes | 1,899 (11.8%) | 46 (9.4%) |  |
| Rheumatic Disease | No | 15,765 (97.7%) | 481 (98.8%) | 0.11<sup>a</sup> |
|  | Yes | 374 (2.3%) | 6 (1.2%) |  |
| Liver Disease, mild | No | 14,938 (92.6%) | 441 (90.6%) | 0.098<sup>a</sup> |
|  | Yes | 1,201 (7.4%) | 46 (9.4%) |  |
| Liver Disease, severe | No | 16,005 (99.2%) | 486 (99.8%) | 0.13<sup>a</sup> |
|  | Yes | 134 (0.8%) | 1 (0.2%) |  |
| Hemiplegia | No | 16,073 (99.6%) | 485 (99.6%) | 1.00<sup>a</sup> |
|  | Yes | 66 (0.4%) | 2 (0.4%) |  |
| Chronic Kidney Disease | No | 14,905 (92.4%) | 444 (91.2%) | 0.33<sup>a</sup> |
|  | Yes | 1,234 (7.6%) | 43 (8.8%) |  |
| Lymphoma | No | 15,921 (98.6%) | 483 (99.2%) | 0.32<sup>a</sup> |
|  | Yes | 218 (1.4%) | 4 (0.8%) |  |
| Leukemia | No | 15,889 (98.5%) | 479 (98.4%) | 0.87<sup>a</sup> |
|  | Yes | 250 (1.5%) | 8 (1.6%) |  |
| Solid Cancer | No | 13,964 (86.5%) | 454 (93.2%) | <0.001<sup>a</sup> |
|  | Yes | 2,175 (13.5%) | 33 (6.8%) |  |
| Metastatic Cancer | No | 15,699 (97.3%) | 479 (98.4%) | 0.15<sup>a</sup> |
|  | Yes | 440 (2.7%) | 8 (1.6%) |  |
| Body Mass Index (kg/m<sup>2</sup>) |  | 26.1 (23.1-30.0) | 27.2 (23.5-31.7) | <0.001<sup>b</sup> |
| BMI ≥ 30 kg/m<sup>2</sup> | No | 12,011 (74.4%) | 317 (65.1%) | <0.001<sup>a</sup> |
|  | Yes | 3,911 (24.2%) | 163 (33.5%) |  |
|  | Missing | 217 (1.3%) | 7 (1.4%) |  |
| Mental Health Diagnosis | No | 10,505 (65.1%) | 320 (65.7%) | 0.78<sup>a</sup> |
|  | Yes | 5,634 (34.9%) | 167 (34.3%) |  |
| Solid Organ Transplant | No | 15,887 (98.4%) | 477 (97.9%) | 0.39<sup>a</sup> |
|  | Yes | 252 (1.6%) | 10 (2.1%) |  |
| Bone Marrow Transplant | No | 16,040 (99.4%) | 484 (99.4%) | 0.99<sup>a</sup> |
|  | Yes | 99 (0.6%) | 3 (0.6%) |  |
| Charlson Comorbidity Index |  | 2 (1-4) | 1 (0-3) | <0.001<sup>b</sup> |

a. Pearson's chi-squared; b. Wilcoxon rank-sum

**Supplementary Table 4:** Patient characteristics by HIV status for in care at UCSD, tested for COVID-19 and diagnosed with COVID-19

|  | In care at UCSD |  | Tested for COVID-19 |  | COVID-19 diagnosis |  |
| --- | --- | --- | --- | --- | --- | --- |
|  | Non-HIV | HIV | Non-HIV | HIV | Non-HIV | HIV |
| Number of people, n (%) | 58,302 | 4,017 | 15,401 (26%) | 1,225 (30%) | 399 (2%) | 88 (7%) |
| **Age, years (median, IQR)** | 57 (39-71) | 51 (40-59)*** | 61 (44-72) | 54 (43-60)*** | 50 (34-64) | 45 (36-53)*** |
| 18-35 years, n (%) | 10,582 (18%) | 578 (14%) | 2,067 (13%) | 142 (11%) | 101 (25%) | 18 (20%) |
| 35-60 years, n (%) | 20,770 (36%) | 2,506 (62%) | 5,158 (33%) | 749 (61%) | 168 (42%) | 57 (65%) |
| >=60 years, n (%) | 26,950 (46%) | 933 (23%) | 8,176 (53%) | 334 (27%) | 130 (32%) | 13 (15%) |
| **Gender, n (%)** |  | *** |  | *** |  | *** |
| Male | 24,628 (42%) | 3,455 (86%) | 6,406 (42%) | 1,058 (86%) | 181 (45%) | 70 (80%) |
| Female | 33,671 (58%) | 562 (14%) | 8,994 (58%) | 167 (14%) | 217 (54%) | 18 (20%) |
| Missing | 3 (0%) | 0 (0%) | 1 (0%) | 0 (0%) | 1 (0%) | 0 (0%) |
| **Race/Ethnicity, n (%)** |  | *** |  | *** |  | *** |
| White | 36,435 (62%) | 1,871 (46%) | 10,312 (67%) | 596 (49%) | 207 (52%) | 27 (31%) |
| Non-White | 21,867 (38%) | 2,146 (53%) | 5,089 (33%) | 629 (51%) | 192 (48%) | 61 (69%) |
| **Social History** |  |  |  |  |  |  |
| Active Smoking, n (%) | 2,814 (5%) | 1,502 (26%)*** | 612 (4%) | 300 (24%)*** | 10 (2%) | 10 (11%)*** |
| Active Drinking, n (%) | 33,342 (57%) | 1,568 (39%)*** | 8,834 (57%) | 491 (40%)*** | 214 (54%) | 51 (58%) |
| Active Drug Use, n (%) | 4,027 (7%) | 1,052 (26%)*** | 1,205 (8%) | 367 (30%)*** | 37 (9%) | 22 (25%)*** |
| Homeless, n (%) | 80 (0%) | 156 (4%)*** | 80 (0%) | 156 (13%)*** | 3 (1%) | 7 (8%)*** |
| **Comorbidities** |  |  |  |  |  |  |
| Hypertension, n (%) | 14,403 (25%) | 1,088 (27%)*** | 4,493 (29%) | 401 (33%)** | 98 (24%) | 22 (25%) |
| Coronary Artery Disease, n (%) | 3,427 (6%) | 207 (5%) | 1,300 (8%) | 89 (7%) | 29 (7%) | 4 (4%) |
| Congestive heart failure, n (%) | 1,878 (3%) | 133 (3%) | 743 (5%) | 59 (5%) | 17 (4%) | 0 (0%) |
| Any diabetes, n (%) | 5,206 (9%) | 374 (9%) | 1,662 (11%) | 148 (12%) | 52 (13%) | 8 (9%) |
| Chronic pulmonary disease, n (%) | 4,856 (8%) | 515 (13%)*** | 1,743 (11%) | 202 (16%)*** | 35 (9%) | 11 (12%) |
| Rheumatic disease, n (%) | 964 (2%) | 12 (0%)*** | 375 (2%) | 5 (0%)*** | 5 (1%) | 1 (1%) |
| Liver Disease, n (%) | 2,905 (5%) | 525 (13%)*** | 1,059 (7%) | 212 (17%)*** | 28 (7%) | 18 (20%)*** |
| Renal disease, n (%) | 3,213 (6%) | 297 (7%)*** | 1,147 (7%) | 130 (11%)*** | 35 (9%) | 8 (9%) |
| Any Cancer, n (%) | 6,158 (10%) | 399 (10%) | 2,482 (16%) | 162 (13%)** | 37 (9%) | 7 (8%) |
| Mental health, n (%) | 15,951 (27%) | 2,541 (63%*** | 4,934 (32%) | 867 (71%*** | 107 (27%) | 60 (68%*** |
| BMI $\geq 30$ kg/m<sup>2</sup>, n (%) | 13,431 (23%) | 995 (25%)** | 3,741 (24%) | 333 (27%) | 132 (33%) | 31 (35%) |
| Solid organ transplant, n (%) | 500 (1%) | 15 (0%*** | 253 (2%) | 9 (1%)* | 10 (2%) | 0 (0%) |
| Charlson comorbidity index, median (IQR) | 2 (0-4) | 1 (0-3)*** | 2 (1-4) | 2 (0-3)*** | 1 (0-3) | 1 (0-2) |
| **HIV Risk Factors, n (%)** |  |  |  |  |  |  |
| MSM |  |  | - | 791 (64%) | - | 56 (64%) |
| Heterosexual |  |  | - | 264 (22%) | - | 25 (28%) |
| Hemophilia |  |  | - | 3 (0%) | - | 1 (1%) |
| MSM+IDU |  |  | - | 65 (5%) | - | 3 (3%) |
| Heterosexual + IDU |  |  | - | 30 (2%) | - | 2 (2%) |
| Other |  |  | - | 4 (0%) | - | 0 (0%) |
| Missing |  |  | - | 68 (6%) | - | 1 (1%) |
| **CD4 count (median, IQR)** |  |  | - | 634 (405-881) | - | 638 (406-1014) |
| <200, n (%) |  |  | - | 89 (7%) | - | 4 (4%) |
| $\geq 200$, n (%) | | | - | 1037 (85%) | - | 80 (91%) |
| Missing |  |  | - | 99 (8%) | - | 4 (4%) |
| **Log(10) VL (median, IQR)** |  |  | - | 1.30 (1.30-1.33) | - | 1.30 (1.30-1.30) |
| $\leq 50$, n (%) | | | - | 984 (80%) | - | 74 (84%) |
| >50, n (%) |  |  | - | 169 (14%) | - | 10 (11%) |
| Missing, n (%) |  |  | - | 72 (6%) | - | 4 (4%) |
| **ART** |  |  | - | 1204 (98%) | - | 88 (100%) |
| INSTI |  |  | - | 1057 (86%) | - | 78 (88%) |
| PI |  |  | - | 163 (13%) | - | 11 (12%) |
| NNRTI |  |  | - | 163 (13%) | - | 11 (12%) |
| Tenofovir |  |  | - | 951 (78%) | - | 66 (75%) |

\*Indicates p<0.05 for difference between PWH and non-PWH groups via pooled t-test and Wilcoxon rank-sum test for continuous variables and Fisher's exact test for categorical variables
\*\*Indicates p< 0.01 and \*\*\* indicates< 0.0001 for the same comparisons

**Supplementary Figure 1:**
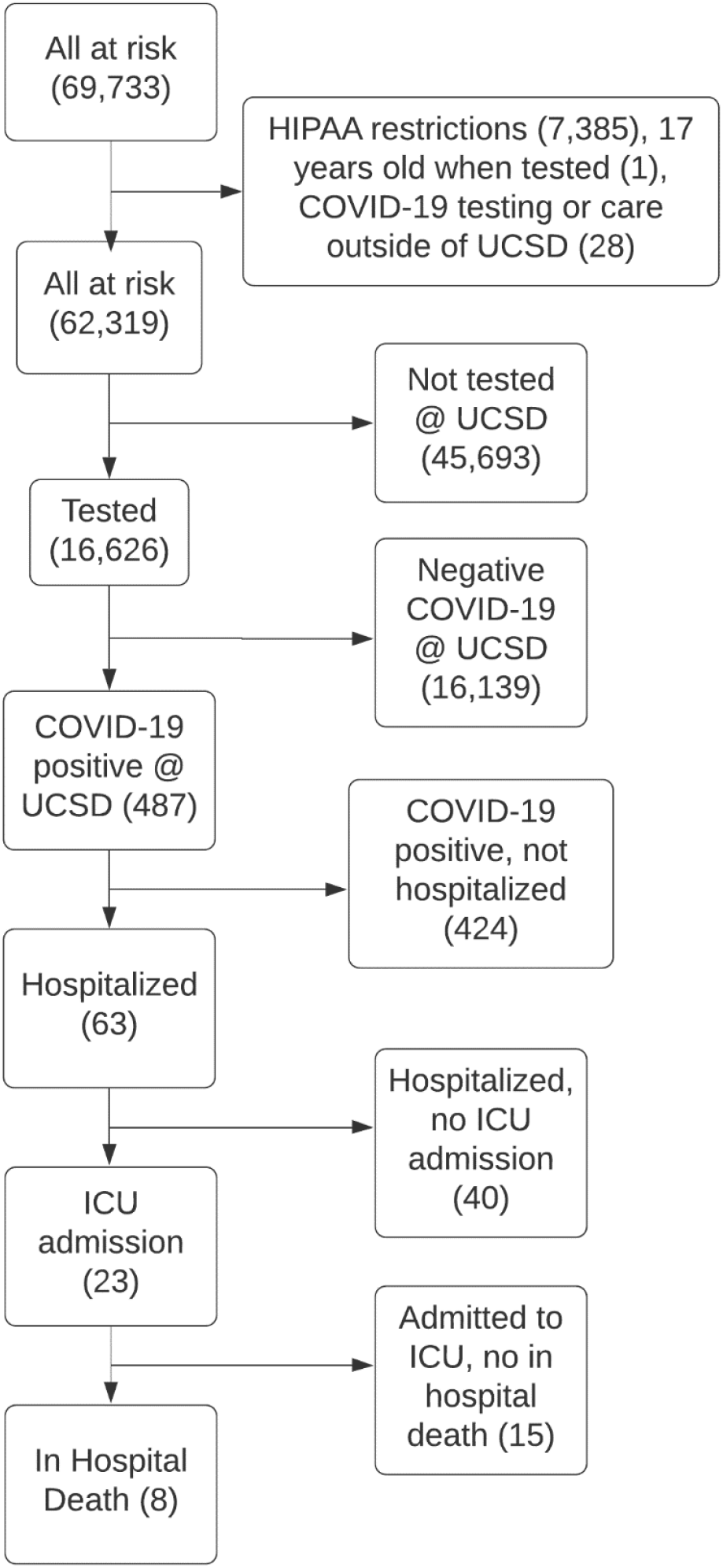
Flow diagram for patients included in analysis

## References

1. Friedman MR, Stall R, Silvestre AJ, et al. Effects of syndemics on HIV viral load and medication adherence in the multicentre AIDS cohort study. AIDS 2015; 29:1087–1096.

2. Bhaskaran K, Rentsch CT, MacKenna B, et al. HIV infection and COVID-19 death: a population-based cohort analysis of UK primary care data and linked national death registrations within the OpenSAFELY platform. Lancet HIV 2021; 8:e24–e32.

3. Western Cape Department of Health in collaboration with the National Institute for Communicable Diseases, South Africa, Boulle A, Davies M-A, et al. Risk Factors for Coronavirus Disease 2019 (COVID-19) Death in a Population Cohort Study from the Western Cape Province, South Africa. Clin Infect Dis 2020; :ciaa1198.

4. del Amo J, Polo R, Moreno S, et al. Incidence and Severity of COVID-19 in HIV-Positive Persons Receiving Antiretroviral Therapy: A Cohort Study. Ann Intern Med 2020; 173:536–541.

5. Huang J, Xie N, Hu X, et al. Epidemiological, Virological and Serological Features of Coronavirus Disease 2019 (COVID-19) Cases in People Living With Human Immunodeficiency Virus in Wuhan: A Population-based Cohort Study. Clin Infect Dis 2021; 73:e2086–e2094.

6. Hadi YB, Naqvi SFZ, Kupec JT, Sarwari AR. Characteristics and outcomes of COVID-19 in patients with HIV: a multicentre research network study. AIDS 2020; 34:F3–F8.

7. Dandachi D, Geiger G, Montgomery MW, et al. Characteristics, Comorbidities, and Outcomes in a Multicenter Registry of Patients With Human Immunodeficiency Virus and Coronavirus Disease 2019. Clin Infect Dis Off Publ Infect Dis Soc Am 2021; 73:e1964–e1972.

8. Karmen-Tuohy S, Carlucci PM, Zervou FN, et al. Outcomes Among HIV-Positive Patients Hospitalized With. J Acquir Immune Defic Syndr 2020; 85:5.

9. Sachdev D, Mara E, Hsu L, et al. COVID-19 Susceptibility and Outcomes Among People Living With HIV in San Francisco. JAIDS J Acquir Immune Defic Syndr 2021; 86:19–21.

10. Gervasoni C, Meraviglia P, Riva A, et al. Clinical features and outcomes of HIV patients with coronavirus disease 2019. Clin Infect Dis 2020; :ciaa579.

11. Park LS, Rentsch CT, Sigel K, et al. COVID-19 in the largest US HIV cohort. 2020.

12. Braunstein SL, Lazar R, Wahnich A, Daskalakis DC, Blackstock OJ. Coronavirus Disease 2019 (COVID-19) Infection Among People With Human Immunodeficiency Virus in New York City: A Population-Level Analysis of Linked Surveillance Data. Clin Infect Dis 2021; 72:e1021–e1029.

13. Tesoriero JM, Swain C-AE, Pierce JL, et al. COVID-19 Outcomes Among Persons Living With or Without Diagnosed HIV Infection in New York State. JAMA Netw Open 2021; 4:e2037069.

14. Durstenfeld MS, Sun K, Ma Y, et al. Impact of HIV on COVID-19 outcomes among hospitalized adults in the United States. 2021. Available at: https://theprogramme.ias2021.org/Abstract/Abstract/2377.

15. Bertagnolio S, Thwin SS, Silva R, et al. Clinical characteristics and prognostic factors in people living with HIV hospitalized with COVID-19: findings from the WHO Global Clinical Platform. 2021. Available at: https://theprogramme.ias2021.org/Abstract/Abstract/2498.

16. Charlson ME, Pompei P, Ales KL, MacKenzie CR. A new method of classifying prognostic comorbidity in longitudinal studies: development and validation. J Chronic Dis 1987; 40:373–383.

17. Linton N, Kobayashi T, Yang Y, et al. Incubation Period and Other Epidemiological Characteristics of 2019 Novel Coronavirus Infections with Right Truncation: A Statistical Analysis of Publicly Available Case Data. J Clin Med 2020; 9:538.

18. Lauer SA, Grantz KH, Bi Q, et al. The Incubation Period of Coronavirus Disease 2019 (COVID-19) From Publicly Reported Confirmed Cases: Estimation and Application. Ann Intern Med 2020; 172:577–582.

19. Griffith GJ, Morris TT, Tudball MJ, et al. Collider bias undermines our understanding of COVID-19 disease risk and severity. Nat Commun 2020; 11:5749.

20. Seaman SR, White IR. Review of inverse probability weighting for dealing with missing data. Stat Methods Med Res 2013; 22:278–295.

21. Thoemmes F, Ong AD. A Primer on Inverse Probability of Treatment Weighting and Marginal Structural Models. Emerg Adulthood 2016; 4:40–59.

22. Haneuse S, VanderWeele TJ, Arterburn D. Using the E-Value to Assess the Potential Effect of Unmeasured Confounding in Observational Studies. JAMA 2019; 321:602–603.

23. VanderWeele TJ, Ding P. Sensitivity Analysis in Observational Research: Introducing the E-Value. Ann Intern Med 2017; 167:268.

24. Chang JJ, Bruxvoort K, Chen LH, Akhavan B, Rodriguez J, Hechter RC. Brief Report: COVID-19 Testing, Characteristics, and Outcomes Among People Living With HIV in an Integrated Health System. J Acquir Immune Defic Syndr 1999 2021; 88:1–5.

25. Díez C, Del Romero-Raposo J, Mican R, et al. COVID-19 in hospitalized HIV-positive and HIV-negative patients: A matched study. HIV Med 2021; 22:867–876.

26. Yendewa GA, Perez JA, Schlick K, Tribout H, McComsey GA. Clinical Features and Outcomes of Coronavirus Disease 2019 Among People With Human Immunodeficiency Virus in the United States: A Multicenter Study From a Large Global Health Research Network (TriNetX). Open Forum Infect Dis 2021; 8:ofab272.

27. Spinelli MA, Brown LB, Glidden DV, et al. SARS-CoV-2 incidence, testing rates, and severe COVID-19 outcomes among people with and without HIV. AIDS Lond Engl 2021; 35:2545–2547.

28. Inciarte A, Gonzalez-Cordon A, Rojas J, et al. Clinical characteristics, risk factors, and incidence of symptomatic coronavirus disease 2019 in a large cohort of adults living with HIV: a single-center, prospective observational study. AIDS Lond Engl 2020; 34:1775–1780.

29. Laracy J, Zucker J, Castor D, et al. HIV-1 Infection Does Not Change Disease Course or Inflammatory Pattern of SARS-CoV-2-Infected Patients Presenting at a Large Urban Medical Center in New York City. Open Forum Infect Dis 2021; 8:ofab029.

30. Sigel K, Swartz T, Golden E, et al. Coronavirus 2019 and People Living With Human Immunodeficiency Virus: Outcomes for Hospitalized Patients in New York City. Clin Infect Dis Off Publ Infect Dis Soc Am 2020; 71:2933–2938.

31. Nomah DK, Reyes-Urueña J, Díaz Y, et al. Sociodemographic, clinical, and immunological factors associated with SARS-CoV-2 diagnosis and severe COVID-19 outcomes in people living with HIV: a retrospective cohort study. Lancet HIV 2021; 8:e701–e710.

32. Shapiro AE, Bender Ignacio RA, Whitney BM, et al. Factors associated with severity of COVID-19 disease in a multicenter cohort of people with HIV in the United States, March-December 2020. MedRxiv Prepr Serv Health Sci 2021; :2021.10.15.21265063.

33. Kirk G. COVID-19 Disease Severity among People with HIV Infection or Solid Organ Transplant in the United States: A Nationally-representative, Multicenter, Observational Cohort Study. :34.

